# Multilocus drug resistance mutation analysis in 148 HBV patients in northern Henan Province of China

**DOI:** 10.1101/2021.05.14.21257253

**Authors:** Zexin Zhou, Minglei Chen, Penghua Li, Junzheng Yang

## Abstract

**Objectives:** To analyze the genotypes and reverse transcriptase mutations gene sites in hepatitis B (HBV) patients in northern Henan Province from 2016 to 2018, which treated by tenofovir dipivoxil(TDF), Lbivudine(LDT), adefovir(ADV), lamivudine (LAM) and entecavir(ETV) analogues, to provide evidences for rational drug use and antiviral therapy for HBV patients.

**Methods:** HBV patients’ serum DNA was extracted, specific primers were designed for YMDD mutation sites in conservative regions; DNA sequencing of the PCR products was carried out by PCR amplification combined with Dideoxygenation termination sequencing(DDT), and then analyzed genotype and reverse transcriptase mutations gene sites of HBV patients.

**Results:** We collected 148 samples of HBV patients in northern Henan Province from 2016 to 2018; 97.97% of HBV patients were genotype C and 2.03% were genotype B; there are 46 cases of LAM and FTC resistance mutation, respectively; 28 cases of LDT resistance mutation, 10 cases of ADV resistance mutation, 8 cases of ETV resistance mutation, and 1 case of TDF resistance mutation were detected; the mutation rates of drug resistance was 43.45% in HBV patients with genotype C and 33.33% in HBV patients with genotype B. There was no significant difference in the distribution rate between the two genotypes (P>0.05). The most common mutations types were M204M|V and M204M|I. The M204M|V mutation type often occurred in combination of M204M|V+L180M|L+L173V|L and M204M|V+L180M|L and M204M|V+ L180M|L+V173V|M, while M204M|I mutation type was often occurred alone, M204M|V+L180M|L combination appeared alone or in combination. N236+A181 base substitution was the dominant site in ADV resistance. ETV resistance mutation was based on LAM and ETV resistance, mainly were T184 and S202 base substitution; and LDT resistance mutation was M204M|I.

**Conclusion:** In this article, PCR and DDT DNA sequencing were used to detect multisite resistance mutations of HBV reverse transcriptase gene in HBV patients. It was found that most of the genotypes of HBV patients in northern Henan Province were genotype C, the combination of drug resistance mutations was complex, and the mutation rates of multidrug resistance genes was high. There was no significant difference in the distribution rate of HBV genotypes among the detected patients, those data may provide evidences for the detection and confirmation of HBV resistance and rational drug use in HBV patients in the region.

## 1. Introduction

HBV is one of the most infectious diseases in China, which is the main factor of liver cirrhosis, liver cancer and liver failure. It has been found that the high HBV DNA concentration in chronic HBV patients’ blood is often related to high risk of liver failure and liver cancer [2, 3], The low level of HBV DNA in blood or in undetected state can greatly reduce the risk of mortality in liver cancer patient or liver failure patient, so it is particularly important to observe and control the HBV DNA concentration in HBV patients [4-6]. In recent years, due to tenofovir [7, 8] Nucleoside (acid) analogues such as tenofovir discoprox(TDF), libivudine(LDT), adefovir(ADV), lamivudine (LAM) [9], Emtricitabine(FTC) [10], entecavir (ETV) [11] and other nucleosides (acids) have been found, and have been widely used in HBV treatment. The target of nucleoside (acid) analogues is HBV polymerase/reverse transcriptase (RT), and drug resistance mutation often occurs in the sub-region, and it is divided into primary and secondary self compensating drug resistance mutations. When HBV is under drug selection pressure, it can induce the virus strain with drug resistance mutation to be amplified selectively or RT gene can produce adaptive drug resistance mutation. Because of the increase of the type of anti-HBV nucleoside (acid) analogues and the extension of the use time, it is impossible to meet the clinical needs only for detecting the lamivudine resistant mutation site YVDD/YIDD(rtM204V/I).

At present, it is known that TDF resistance mutation site is A194T in RT gene region, LDT resistance mutation site is M204I, ADV resistance mutation site are A181V/T and N236T, LAM and FTC resistance mutation site are V173I, I180M and M204I/V, ETV resistance mutations are I169T, I180M, T184A /T184G/T184S/T184I /T184L/T184F, S202G/I, M204V and M250V/L/I. The drug-resistant mutant sites are still emerging. Therefore, detection of all nucleoside (acid) analogues resistance mutations and their change types in HBV RT gene region is of great clinical significance for rational drug use in the HBV treatment. Here, we detected and analyzed the drug-resistant mutation sites of HBV RT gene in 148 chronic HBV patients from northern Henan province of China from 2016 to 2018 by PCR amplification method combined with DDT method, in order to provide the basis for clinical medication and monitoring of HBV patients.

## Materials and methods

### 1. Materials

148 serum samples from HBV patients were collected for detection from 2016 to 2018. The HBV viral load in the serum of patients was 103∼108 copies/mL, and they were diagnosed as chronic hepatitis B. the diagnosis met the diagnostic criteria of chronic toxic hepatitis in the “viral hepatitis prevention and treatment plan” revised in 2000.

### 2. Methods

#### 2.1 HBV RT gene amplification and DNA sequencing

the virus DNA was extracted by the virus nucleic acid extraction kit produced by Sun Yat-sen University Da’an gene Co., Ltd. RT gene region was amplified by PCR sequencing method. PCR products were purified by uniq-10 column PCR product purification kit from Shanghai Shenggong Company. After PCR products were purified, DNA sequence was determined by ABI3130 gene sequencing machine.

#### 2.2 HBV DNA load determination

real time quantitative PCR was used for HBV DNA load determination, the reagent was produced by Shanghai Fuxing company, and the detection limitation was 500 copies/ml.

#### 2.3 Gene mutation analysis

the sequence sites of A194, V173, L180, A181, T184, S202, M204, I169 and M250 were analyzed. There were two peaks at the same site in the sequence, the lower peak reached 25% of the higher peak, which indicated that there was nucleotide coexistence.

### 3. Statistical analysis

χ^2^ test was used to test the difference between the two groups, P < 0.05 means significant difference.

### 4. Results

#### 4.1 Information of HBV patients

The average age was 39.5±2.5 years old in 148 cases of HBV patients in Northern Henan (the minimum age was 13±2 years old, the maximum age was 66±5 years old); 104 cases were male, the average age was 40.5±2.5 years old; 44 cases were female, the average age was 43.5±2.5 years old.

#### 4.2 Genotyping

The results showed that there were 145 cases (97.97%) of HBV C genotype patients, 3 cases (2.03%) of HBV B genotype patients, The mutation rate of drug resistance was 43.45% in genotype C patients and 33.33% in genotype B patients. There was no significant difference between mutation rate of drug resistance in the two genotypes (P > 0.05).

#### 4.3 Detection results of drug resistance mutation in 148 HBV patients

148 cases of HBV patients were collected and RT gene sequence, results showed that drug resistance mutation sites of ETV were I169T (1 case, 0.95%), T184I (2 cases, 1.90%), S202G (2 cases, 1.90%) and T184I (1 case, 0.95%), and no mutation sites of T184A, T184F and T184G were found; the drug resistance mutation sites of ADV were A181T (8 cases, 7.6%). It should be pointed out that LAM and other drug-resistant nucleoside drugs often have the same mutation sites, M204I mutation could occur with LDT/LAM/FTC/ETV combination(18 cases, 17.14%), M204V mutation occur with LAM/FTC/ETV combination (17 cases, 16.19%), L180M mutation occur with LAM/FTC/ETV combination (15 cases, 14.29%) and V137I mutation occur with LAM/FTC combination (7 cases, 6.67%). Data are shown in Table 2.

**Table 1.** Mutation rate of drug resistance between genotype C and B in 148 HBV patients

| Genotype | case | Of the total positive cases | Cases of drug resistance | Mutation rate of drug resistance |
| --- | --- | --- | --- | --- |
| genotype C | 145 | 97.97% | 63 | 43.45% |
| genotype B | 3 | 2.03% | 1 | 33.33% |

**Table 2.** Types, mutation sites and detection rate of drug-resistant nucleoside drugs

| Mutation sites | case | Positive rate | nucleoside drugs |
| --- | --- | --- | --- |
| I169T | 1 | 0.95% | ETV |
| I169L | 1 | 0.95% | undefined |
| I169N | 1 | 0.95% | undefined |
| V173L | 7 | 6.67% | LAM/ FTC |
| V173G | 1 | 0.95% | undefined |
| V173M | 1 | 0.95% | undefined |
| L180M | 15 | 14.29% | LAM/ FTC/ ETV |
| A181T | 8 | 7.62% | ADV |
| A181V | 6 | 5.71% | ADV |
| A181I | 2 | 1.90% | undefined |
| T184I | 2 | 1.90% | ETV |
| T184L | 1 | 0.95% | ETV |
| T184A | 0 | 0.00% | ETV |
| T184F | 0 | 0.00% | ETV |
| T184G | 0 | 0.00% | ETV |
| A194 | 0 | 0.00% | TDF |
| S202G | 2 | 1.90% | ETV |
| M204I | 18 | 17.14% | LDT/LAM/FTC/ETV |
| M204V | 17 | 16.19% | LAM/FTC/ETV |
| N236T | 8 | 7.62% | ADV |
| M250V | 0 | 0.00% | ETV |
| M250L | 0 | 0.00% | ETV |
| M250I | 0 | 0.00% | ETV |

#### 4.4. Drug resistance mutations and major mutation combinations of nucleoside drugs in 148 cases of HBV patients

According to the sequencing results from nucleoside drugs, the most drug-resistant mutations were found in LAM (31 cases) and FTC (31 cases), accounting for 25.62% of the total number of drug-resistant mutations; the major mutation site was M204I (9 cases), accounting for 7.44% of the total drug-resistant mutations, followed by L180M+M204V (8 cases), accounting for 8% of the total number of drug-resistant mutations, V173L+L180M+M204V (6 cases, accounting for 4.96% of the total drug-resistant mutations); ADV resistant mutations were 23 cases, and the combination of gene mutation site was N236T (8 cases, accounting for 6.61% of the total drug-resistant mutations), followed by A181T (7 cases, accounting for 5.79% of the total drug-resistant mutations), A181V (4 cases, accounting for 3.31% of the total drug-resistant mutations). A181V+N236T (2 cases, accounting for 1.65% of the total drug-resistant mutations) and ETV resistant mutations were 18 cases, L181M+M204V (8 cases, accounting for 6.61% of the total drug-resistant mutations), followed by L181M+M204I (4 cases, accounting for 3.31% of the total drug-resistant mutations), L181M+M204V+S202G and L181M+M204V+T184I (2 cases, accounting for 1.65% of the total drug-resistant mutation, respectively), The data were shown in Table 3.

**Table 3.** Major mutation sites of drug-resistant nucleoside drugs and the proportion of drug-resistant mutations in 148 HBV patients

| Nucleoside drugs | Case | Positive rate in 148 HBV patients | Mutation site | case | The proportion of drug-resistant mutations in nucleoside drugs | The proportion in total drug-resistant mutations |
| --- | --- | --- | --- | --- | --- | --- |
| LDT | 17 | 14.05% | M204I | 17 | 100.00% | 14.05% |
| ADV | 23 | 19.01% | A181V | 4 | 17.39% | 3.31% |
|  |  |  | A181T | 7 | 30.43% | 5.79% |
|  |  |  | N236T | 8 | 34.78% | 6.61% |
|  |  |  | A181T+N236T | 1 | 4.35% | 0.83% |
|  |  |  | A181V+N236T | 2 | 8.70% | 1.65% |
|  |  |  | A181I+N236T | 1 | 4.35% | 0.83% |
| LAM | 31 | 25.62% | M204V | 2 | 6.45% | 1.65% |
|  |  |  | M204I | 9 | 32.26% | 7.44% |
|  |  |  | L180M+M204V | 8 | 25.81% | 6.61% |
|  |  |  | L180M+M204I | 4 | 12.90% | 3.31% |
|  |  |  | V173L+L180M+M204I | 1 | 3.23% | 0.83% |
|  |  |  | V173L+L180M+M204V | 6 | 19.35% | 4.96% |
|  |  |  | V173G+M204I | 1 | 3.23% | 0.83% |
| FTC | 31 | 25.62% | M204V | 2 | 6.45% | 1.65% |
|  |  |  | M204I | 9 | 32.26% | 7.44% |
|  |  |  | L180M+M204V | 8 | 25.81% | 6.61% |
|  |  |  | L180M+M204I | 4 | 12.90% | 3.31% |
|  |  |  | V173L+L180M+M204I | 1 | 3.23% | 0.83% |
|  |  |  | V173L+L180M+M204V | 6 | 19.35% | 4.96% |
|  |  |  | V173G+M204I | 1 | 3.23% | 0.83% |
| ETV | 18 | 14.88% | L180M+M204V | 8 | 44.44% | 6.61% |
|  |  |  | L180M+M204I | 4 | 22.22% | 3.31% |
|  |  |  | L180M+M204V+S202G | 2 | 11.11% | 1.65% |
|  |  |  | L180M+M204V+T184L | 1 | 5.56% | 0.83% |
|  |  |  | L180M+M204V+T184I | 2 | 11.11% | 1.65% |
|  |  |  | L180M+M204V+T184S | 1 | 5.56% | 0.83% |
| TDF | 1 | 0.83% | A194T | 1 | 100% | 0.83% |

### 5. Discussion

In the past, many studies have reported that HBV drug-resistant mutations occur in the process of HBV treatment. Accurate and real-time monitoring and observation of the occurrence characteristics of HBV drug-resistant mutations have important clinical significance for the treatment of HBV and the reduction of liver cancer or liver failure caused by HBV [12]. In this paper, we reported 148 cases of HBV multi site drug resistance related mutations in Northern Henan of China, and sorted out the relevant results. We found that HBV genotype in Northern Henan was mainly type C, accounting for 97.97% of the total detected HBV patients, which was consistent with most of the related reports. Henan is in the center of China, and HBV genotypes in China are mainly divided into type A, B, C, D, while in the north of China Most of them were B and C genotypes. The statistical results showed that no matter genotype B or genotype C, there was no significant difference in the mutation rate of HBV resistance gene, in which genotype C was 43.45%, genotype B was 33.33%; it needs to be specially pointed out that for HBV patients in Northern Henan, although only 3 cases of genotype B were detected, there was 1 case of HBV resistance mutation, which needs to be paid enough attention.

The results of base mutation detection showed that in 148 cases of HBV patients in Northern Henan Province, different drug-resistant nucleoside drugs had different gene mutations, including M204I mutation (18 cases, 17.14%), M204V mutation (17 cases, 16.19%) and L180M mutation (15 cases, 14.29%), accounting for 17.14%, 16.19% and 14.29% in the total mutations, respectively, These three mutation sites correspond to LDT/LAM/FTC/ETV, LAM/FTC/ETV and LAM/FTC/ETV, respectively. It is suggested that we should avoid the simultaneous use of these nucleoside drugs, especially LAM and FTC.

Based on the analysis of drug-resistant mutations and major mutation combinations of 148 HBV patients in Northern Henan Province, it was found that LAM and FTC had the most drug-resistant mutations (31 cases, respectively), and the major mutation site were M204I, L180M+M204V, V173l+L180M+M204V, in view of this result, we should avoid the use of LAM and FTC, or use the mixed and alternative use of other nucleoside drugs, try to reduce and shorten the dosage and time of LAM and FTC, and reduce the probability of HBV drug resistance mutation; It should be pointed out that the number of drug-resistant mutations of ADV and ETV reached 23 and 18 respectively, which should be paid attention to and used rationally.

It should be pointed out that two HBV patients aged 10-16 had mutations at L108+M204V and M204I sites, respectively, which revealed that HBV drug resistance mutations may be inherited and have a relationship with their own tolerance, which suggest us the importance of rational drug use.

In summary, we analyzed 148 cases of HBV patients of drug resistance mutations in Northern Henan Province from 2016 to 2018. It was found that the main genotype of HBV patients in Northern Henan Province was type C, and the main mutation sites were M204I mutation (18 cases, 17.14%), M204V mutation (17 cases, 16.19%), L180M mutation (15 cases, 14.29%), The number of drug-resistant mutations in LAM and FTC were the most (31 cases each), and the main mutation sites were M204I, L180M+M204V, V173L+L180M+M204V, which suggested that we should pay attention to the rational mixed use of drugs to prevent the occurrence of drug resistance.

## Data Availability

The data used to support the findings of this study are available from the corresponding author upon request

## 6. Acknowledgement

None.

